# Will the COVID-19 pandemic lead to a tsunami of suicides? A Swedish nationwide analysis of historical and 2020 data

**DOI:** 10.1101/2020.12.10.20244699

**Authors:** Christian Rück, David Mataix-Cols, Kinda Malki, Mats Adler, Oskar Flygare, Bo Runeson, Anna Sidorchuk

**Author notes:** Corresponding author: Professor Christian Rück, M46, Karolinska University Hospital Huddinge, SE-14186 Huddinge, Sweden.

## Abstract

**Background:** Various surveys have documented a negative impact of the COVID-19 pandemic on the population’s mental health. There is widespread concern about a surge of suicides, but evidence supporting a link between global pandemics and suicide is very limited. Using historical data from the three major influenza pandemics of the 20^th^ century, and recently released data from the first half of 2020, we aimed to investigate whether an association exists between influenza deaths and suicide deaths.

**Methods:** Annual data on influenza death rates and suicide rates were extracted from the Statistical Yearbook of Sweden from 1910-1978, covering the three 20^th^ century pandemics, and from Statistics Sweden for the period from January to June of each year during 2000-2020. COVID-19 death data were available for the first half of 2020. We implemented non-linear autoregressive distributed lag (NARDL) models to explore if there is a short-term and/or long-term effect of increases and decreases in influenza death rates on suicide rates during 1910-1978. Analyses were done separately for men and women. Descriptive analyses were used for the available 2020 data.

**Findings:** Between 1910-1978, there was no evidence of either short-term or long-term significant associations between influenza death rates and changes in suicides. The same pattern emerged in separate analyses for men and women. Suicide rates in January-June 2020 revealed a slight decrease compared to the corresponding rates in January-June 2019 (relative decrease by −1.2% among men and −12.8% among women).

**Interpretation:** We found no evidence of short or long-term association between influenza death rates and suicide death rates across three 20^th^ century pandemics or during the first six months of 2020 (when the first wave of COVID-19 occurred). Concerns about a substantial increase of suicides may be exaggerated. The media should be cautious when reporting news about suicides during the current pandemic.

## INTRODUCTION

Various international surveys have documented a negative impact of the COVID-19 pandemic on the population’s mental health, with increased levels of psychological stress, psychiatric symptoms, insomnia, and alcohol consumption.[1–4] A report from the UK also indicated an increase in suicidal ideation.[3] Whether these acute impacts will persist long term is currently unknown. The Royal College of Psychiatrists in the UK has warned of a “tsunami of mental illness”, and the World Health Organization as well as the International Academy of Suicide Research have raised concerns of a possible increase in suicide rates.[5–7] Such concerns originate from a combination of known risk factors for suicide, including the impacts of social distancing and disconnectedness, an economic downturn, the decreased access to mental health services, and increased access to lethal means exemplified by an increase in gun purchases in the US.[8] For example, a study of the economic recession in USA 2007-2009 found that for every percentage point increase in the unemployment rate, there was about a 1.6 per cent increase in the suicide rate.[9,10] By contrast, a study failed to find an increase in suicide rates in Sweden during the two most recent economic recessions.[11]

While the concern is widespread, there is currently little evidence to support a clear association between the ongoing COVID-19 outbreak and an increased risk of suicide. A recent review concluded that the quality of the evidence so far on the association between COVID-19 and suicidal behaviour was low and initial reports do not indicate an acute increase of suicides in the first COVID-19 wave.[12,13] Historic US mortality data from the largest pandemic in the 20^th^ century, the Spanish flu, showed that the Spanish flu was associated with an increase in suicides but those effects may have been mitigated by a decline in alcohol consumption.[14] The outbreak of the Severe Acute Respiratory Syndrome (SARS) epidemic in 2003 in Hong Kong was according to two studies associated with an increase of suicide in the population above the age of 65.[15,16] In the first of these two reports, the association was only statistically significant in elderly women.[15] To summarize, at present, our understanding of the effects of pandemics on suicide rates is very limited.

The availability of high-quality historical data on mortality due to influenza and suicide in Sweden provides a unique opportunity to examine this important question. Using historical data from the three major influenza pandemics of the 20^th^ century, we aimed to formally investigate whether an association between influenza deaths and suicide deaths exists. Given the substantial sex differences in suicidal behaviour, we additionally report on associations separately in men and women. We also report preliminary nationwide data from the first half of 2020.

## METHODS

### Data acquisition

Annual data on influenza death rates and suicide rates were estimated based on information from the Statistical Yearbook of Sweden from 1910-1978.[17] Over this period, Sweden was hit by three influenza pandemics which ocurred during different socio-political contexts, namely the Spanish flu (1918-1920, with the first case appearing in Sweden in June 1918), the Asian flu (1957-1958, being first documented in August 1957), and the Hong Kong flu (1968-1969, starting in the autumn of 1968).[18] The yearbooks were produced by Statistics Sweden, a governmental agency responsible for the official statistics in Sweden, with a history of population statistics going back to the 18^th^ century.[17] For each year from 1910 to 1978, we retrieved information on the total population of Sweden, the number of deaths by influenza (if death cause was indicated as “influenza”) and the number of suicides (if death cause was indicated as “suicide”), as well as the corresponding data separately for men and women. Since yearbooks reported data retrospectively for several years prior to the year each book was published, we checked the correctness of retrieved data by comparing yearbooks with overlapping reporting periods. We constructed influenza mortality rates and suicide rates per 100 000 inhabitants for each year by dividing the number of deaths by influenza and, separately, suicide, by the total number of individuals registered in Sweden in a corresponding year and then multiplying by 100 000. The corresponding rates for men and women were constructed likewise. In addition, we collected information on the changes in registration of deaths in Sweden that included the cause of death classification based on the Bertillon criteria (prior to 1931), the new classification introduced in cooperation with statistical authorities from other Nordic countries (1931-1950), the International Classification of Diseases (ICD) Sixth Revision (ICD-6; 1951-1957), ICD-7 (1958-1968), and ICD-8 (1969-1978).[19] To capture a potential effect of changes in classification, we created a series of dummy variables for each corresponding period, but these were only kept in the models if statistically significant.

On November 17^th^, 2020, Statistics Sweden published statistics on the number all-cause and cause specific deaths (for 1997-2020) and the corresponding age-standardized death rates per 100 000 inhabitants for men and women (for 2000-2020; with the total population of Sweden in 2019 as the standard population for both men and women).[20] Importantly, all reported mortality data refer to deaths that occurred during January-June of each year from 1997 to 2020. The reported statistics also included deaths due to COVID-19 that occurred during the first six months of 2020. As the data were made publicly available, we retrieved information on January-June 2000-2020 age-standardized death rates due to influenza (ICD-10 codes: J09 – J11) and suicide and other intentional self-harm (ICD-10 codes: X60 – X84, Y870), and information on January-June 2020 age-standardized death rates due to COVID-19 (ICD-10 codes: U07.1-U07.2), in men and women.

The study protocol was not preregistered. The full dataset used in the analysis of historical data (1910-1978) will be made public upon final publication. The STATA code is available in the online **Supplementary material**. Data on deaths in January-June 1997-2020 are available online at the National Board of Health and Welfare.[20]

### Ethical approval

Since only data already available in the public domain were used, ethical approval was not necessary.

### Statistical Analyses

We used the historical data 1910-1978 that span over three pandemics and focused on exploring a possible asymmetric short-term effect (i.e., instantaneous) and long-term effect (i.e., if distributed over a longer period of time) of influenza death rates on suicides by applying non-linear autoregressive distributed lag (NARDL) models,[21] a technique initially introduced for research in economics. NARDL models split the exposure in partial sum of positive changes (i.e, increases in influenza death rates) and partial sum of negative changes (i.e., decreases in influenza death rates) and explore if an outcome responds differently (i.e., asymmetrically) to an increase and decrease in exposure variable.[21] In other words, NARDL models do not rely on an assumption of symmetrical effects, by which the association of the outcome with a unit of positive change in the exposure is expected to be equal in strength and opposite in direction to the association between the outcome and a unit of negative change in the exposure. We performed the modelling by using the *nardl* command in STATA.

Before the NARDL model is executed, it is important to test that the conditions for the modelling are fulfilled. It starts from testing whether or not the variables are stationary (i.e., their means and variances are constant over time). The advantage of the NARDL modelling is that it can be used regardless of whether the variables are integrated of the order zero, which means that variables are stationary, integrated of the the order one, which indicates non-stationarity, or mixed.[21] We examined the stationarity properties of the variables by using the Augmented Dickey–Fuller unit-root test (ADF) and Kwiatkowski-Phillips-Schmidt-Shin test for stationarity (KPSS) to explore the order of the integration for influenza death rates and suicide rates separately for the total population, men, and women. Previous studies on suicides hypothesized that various suicidogenic factors may interplay and reinforce each other and the logarithmic transformation of suicide time series was suggested.[22,23] We made such transformation by expressing suicide rates in natural logarithm. To select the optimal number of lags (i.e., how far in the past the dependency among measurements is examined) to be used in the NARDL for dependent and independent variables, we applied the *varsoc* command in STATA using the minimal values of Akaike Information Criterion, Schwarz’s Bayesian information criterion (SBIC), and the Hannan and Quinn information criterion information criteria. If information criteria indicated different lag orders, SBIC was used to select optimal lags. We applied the NARDL model to (i) estimate the coefficients and the corresponding 95% confidence intervals for short-term and long-term association of suicide rates with an increase and decrease in influenza death rates; (ii) explore if there is a long-term cointegration between exposure and outcome by using a bounds test (if variables are cointegrated, it means their positive and negative components do not drift far away from each other in the longer term); and (iii) obtain Wald test statistics that specifies whether or not short-term and/or long-term relation between the exposure and outcome is asymmetrical. Model diagnostic tests are described in the **Supplementary material**.

The modelling was performed, first, for the whole period of 1910-1978, and then for the periods of 1918-1956 (from the beginning of the Spanish influenza pandemic to the year before the Asian influenza pandemic), and for 1957-1978 (from the beginning of Asian pandemic to the end of observation period; with this interval also covering the Hong Kong influenza pandemic).

For a descriptive analysis of the data from two recent decades, we graphically visualized age-standardized death rates among men and women for influenza and suicides across January-June 2000-2020, and for COVID-19 in January-June 2020. Additionally, for suicides among men and women, we estimated the relative change in suicide rates reported for the first six months of 2020 to the rates reported for the same months in 2019.

All statistical analyses were performed using STATA version 15.1 (StataCorp LLC, College Station, TX, USA).

## RESULTS

Over the period 1910-1978, influenza death rates fluctuated considerably in Sweden with the highest rates being observed during the Spanish flu (in 1918, 1919, and 1920, the rates were 470.93, 125.55, and 48.32 per 100 000 inhabitants, respectively) (**Figure 1**).

**Figure 1.**
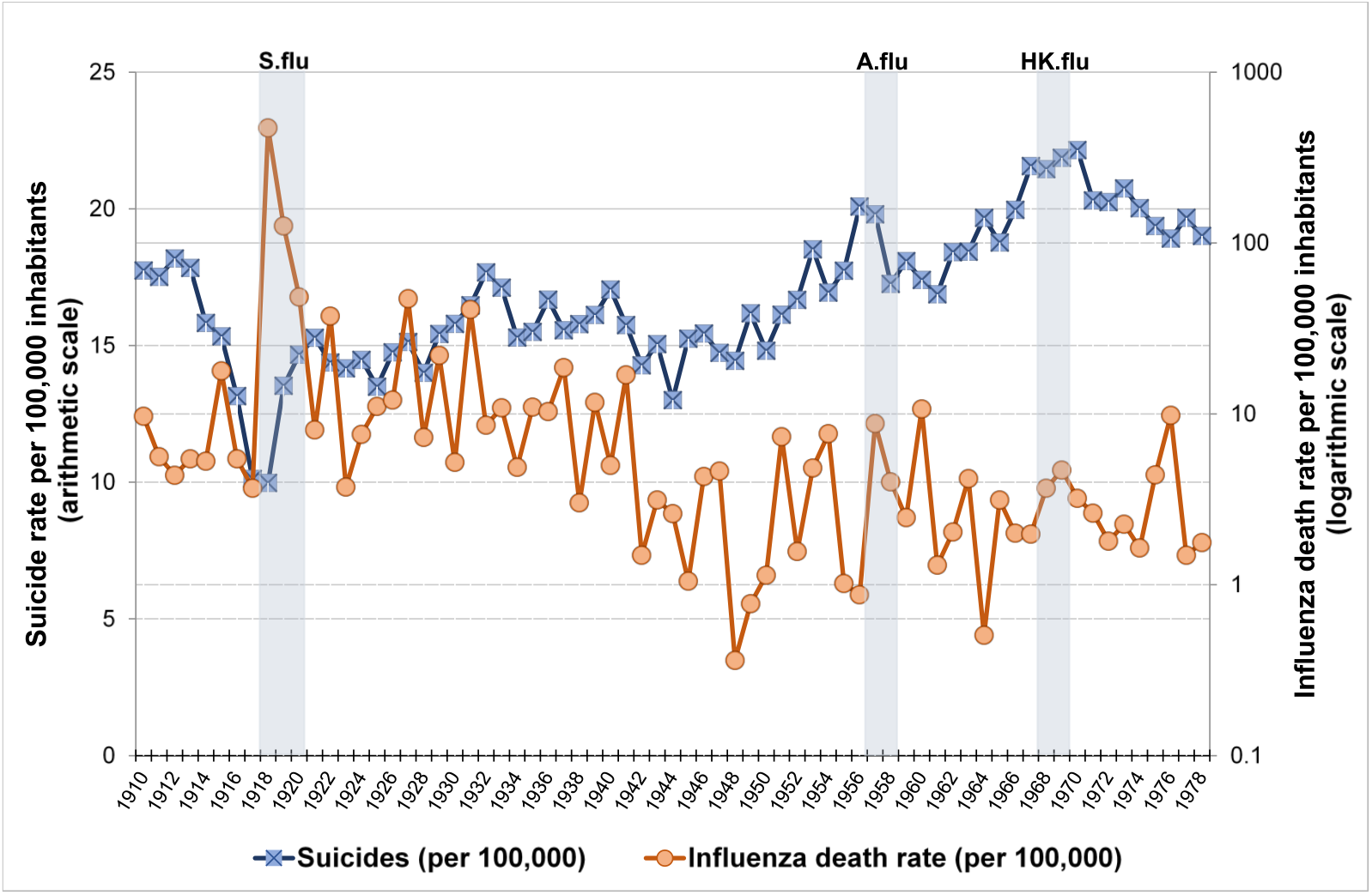
Annual suicide rate (scaled on the left arithmetic Y-axis) and influenza mortality (scaled on the right logarithmic Y-axis) in Sweden in 1910-1978 per 100 000 inhabitants. *Note*: To ease visualization, influenza death rates are reported on the logarithmic scale, while for the analysis, the logarithmic transformation was performed for suicide rates. Grey areas denote the time periods corresponding to the Spanish influenza pandemic (‘S.flu’, 1918-1920), the Asian influenza pandemic (‘A.flu’, 1957-1958), and the Hong Kong influenza pandemic (‘HK.flu’, 1968-1969) in Sweden.

In post-pandemic years, several noticeable peaks in influenza death rates appeared, with the ones in 1922, 1927, 1929, 1931, 1937, and 1941 being particularly high. In the following years, a considerable fluctuation in influenza mortality remained, although the rates during the periods of the Asian flu (in 1957 and 1958: 8.78 and 3.99 per 100 000 inhabitants, respectively) and the Hong Kong flu (in 1968 and 1969: 3.67 and 4.68 per 100 000 inhabitants, respectively) were lower than that in the first half of the century. The influenza death rates were very similar in men and women across the entire observation period (**Figure 2**).

**Figure 2.**
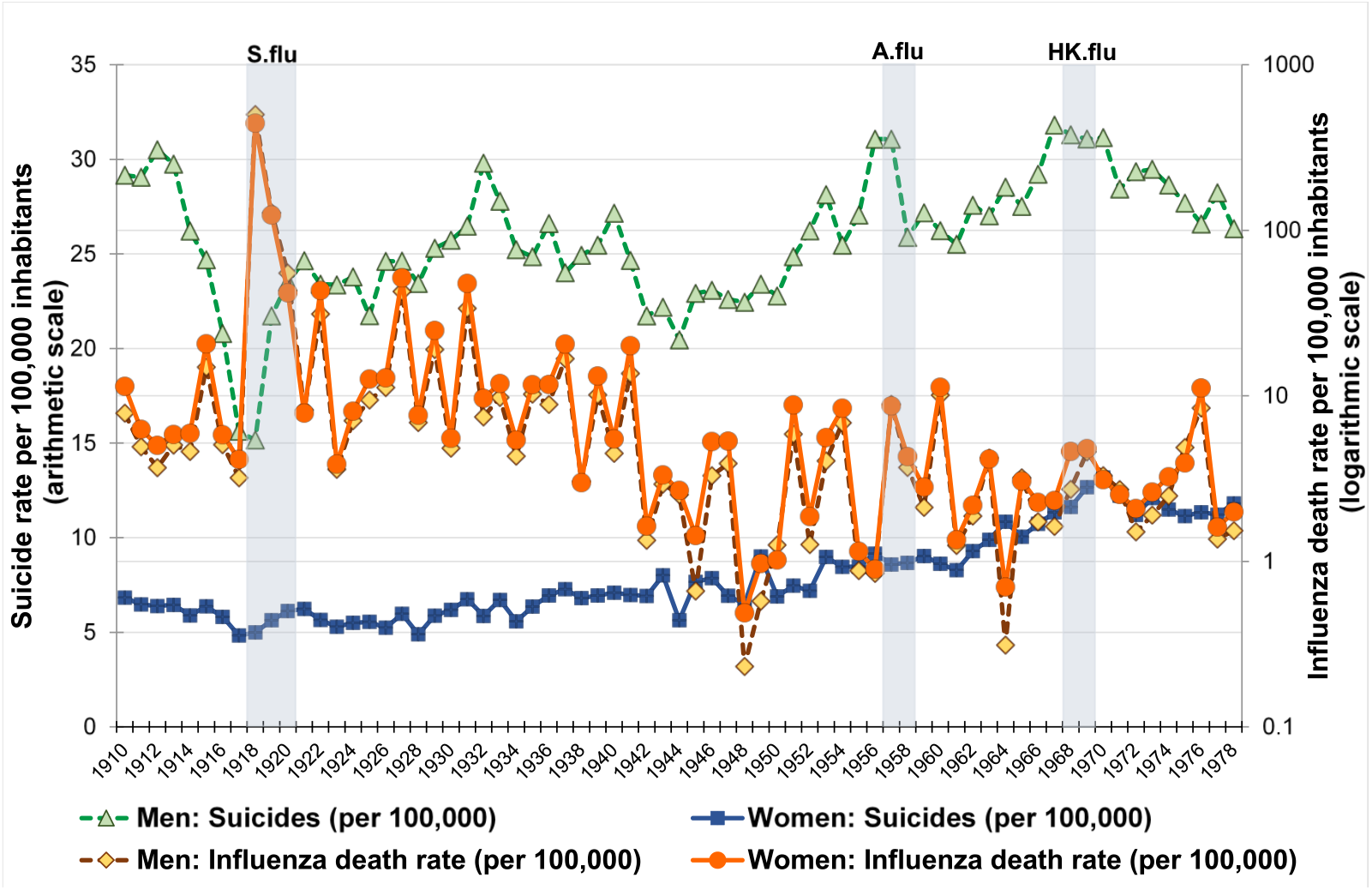
Sex-specific annual suicide rate (scaled on the left arithmetic Y-axis) and influenza mortality (scaled on the right logarithmic Y-axis) in Sweden in 1910-1978 per 100,000 inhabitants of corresponding sex. *Note*: To ease visualization, influenza death rates are reported on the logarithmic scale, while for the analysis, the logarithmic transformation was performed for suicide rates. Grey areas denote the time periods corresponding to the Spanish influenza pandemic (‘S.flu’, 1918-1920), the Asian influenza pandemic (‘A.flu’, 1957-1958), and the Hong Kong influenza pandemic (‘HK.flu’, 1968-1969) in Sweden.

During the same period, a total of 80 058 deaths due to suicide occurred in Sweden (60 713 in men and 19 345 in women). The average suicide rate across 1910-1978 was 16.79 per 100 000 inhabitants (standard deviation [SD] of 2.58), with corresponding rates for men and women as 25.85 (SD=3.37) and 7.91 (SD=2.26) per 100 000, respectively. There was a considerable fluctuation in the suicide rates over time (**Figure 1**). The initial decrease in suicide rates during 1913-1918, with the lowest rate of 9.97 per 100 000 in 1918, was followed by an increase with the highest peak of 22.15 per 100 000 reached in 1970, with a series of intermediate peaks.

Sex-specific suicide rates differed considerably with the rates among men being between twice to over four times higher than that in women (on average, 3.4-times higher) (**Figure 2**). Moreover, suicide rates in men exhibited a sharp dip in 1913-1918, which was first followed by an increase by 1921 and then continued to raise more gradually, although with several distinct peaks and, rarely, dips. Among women, an upward trend in suicide rate was present, although with less fluctuations than that in men.

In our analysis of 1910-1978 data, the first step with the use of the ADF and KPSS test statistics on variables’ stationarity indicated that the logarithmically-transformed suicide rates were integrated of the order one (for all, and men and women separately); whereas the influenza death rates were integrated of the order zero (for all and for men and women separately) (**Supplementary Table 1)**. On this premise, we implemented the NARDL modelling. Dummy variables on changes in death registration did neither appear statistically significant nor affected any parameter estimates in the initial model, and thus were not included in the final models. As reported in **Table 1**, for the observation period of 1910-1978, there were no statistically significant associations (either short-term or long-term) between decreased or increased influenza death rates and suicide rates among the overall population and among men. The corresponding results for women indicated a possible short-term association whereby a decrease in influenza rates seemed to be associated with a borderline decrease in suicides; however, the findings were not supported by the Wald test for asymmetry ([Wald_SR_], i.e. the null hypothesis of Wald_SR_ that an increase and decrease in influenza death rates would symmetrically affect suicide rates was not rejected). This suggests that association was likely due to the effect of other unobserved factors that may influence suicides in women. Full specification of the models is reported in **Supplementary Table 2**. Full specification also includes the results of testing for long-term cointegration between influenza mortality and suicides (as reported by F-statistics for Pesaran/Shin/Smith bounds test [F_PSS]). Long-term cointegration was not found as we were unable to reject the null hypothesis of no cointegration between variables (**Supplementary Table 2**, see footnotes for details on bounds test for cointegration).

**Table 1.** Short-term and long-term relationship of suicide rates with positive and negative changes in influenza death rates among the whole population, men, and women in 1910-1978 in Sweden

|  | Whole population |  | Men |  | Women |  |
| --- | --- | --- | --- | --- | --- | --- |
|  | Coeff. (95% CI) | p-value | Coeff. (95% CI) | p-value | Coeff. (95% CI) | p-value |
| **Short-term coefficients** |  |  |  |  |  |  |
| Influenza + | 0.00002<br>(-0.00036 to 0.00039) | 0.931 | 0.00004<br>(-0.00034 to 0.00041) | 0.854 | -0.00007<br>(-0.00056 to 0.00041) | 0.760 |
| Influenza – | 0.00103<br>(-0.00579 to 0.00785) | 0.764 | -0.00192<br>(-0.01005 to 0.00621) | 0.638 | 0.00780<br>(0.00015 to 0.01544) | 0.046 |
| **Long-term coefficients** |  |  |  |  |  |  |
| Influenza + | -0.00012 | 0.998 | -0.01314 | 0.745 | 0.11254 | 0.538 |
| Influenza – | 0.00211 | 0.962 | 0.01443 | 0.722 | -0.10789 | 0.544 |
| **Model diagnostics** |  |  |  |  |  |  |
| Q-test for autocorrelation, $\chi^2$ | 40.160 | 0.125 | 35.260 | 0.273 | 31.850 | 0.424 |
| Heteroscedasticity, $\chi^2$ | 3.649 | 0.056 | 5.439 | 0.020 | 1.280 | 0.258 |
| Normality, $\chi^2$ | 2.826 | 0.243 | 2.237 | 0.327 | 2.372 | 0.305 |
| RESET, F-statistics | 4.656 | 0.006 | 5.473 | 0.023 | 1.013 | 0.394 |
| CUSUM | Stabl., no str. break | NA | Stabl., no str. break | NA | Stabl., no str. break | NA |
| CUSUMQ | Stabl. | NA | Stabl. | NA | Stabl. | NA |
| Adj. R<sup>2</sup> | 0.257 | NA | 0.300 | NA | 0.374 | NA |
| Wald<sub>SR</sub>, F-statistics | 0.054 | 0.817 | 0.275 | 0.602 | 1.062 | 0.307 |
| Wald<sub>LR</sub>, F-statistics | 1.206 | 0.277 | 1.176 | 0.283 | 0.771 | 0.384 |

*Note:* Signs as “+” and “–” denote the exposure variable being partitioned in positive and negative changes, respectively. Wald test for asymmetry in the short-term (Wald<sub>SR</sub>) and long-term (Wald<sub>LR</sub>) has a null hypothesis of no cointegration. Q-test for autocorrelation reports the results of Portmanteau test for white noise. Heteroscedasticity is measured by Breusch-Pagan/Cook-Weisberg test. Normality is measured by Jarque-Bera test. RESET statistics refers to the regression specification error tests. CUSUM and CUSUMQ refer to the tests of the cumulative sum of recursive residuals and their squares, respectively. “Stabl., no str. break” in the results of CUSUM indicates that the model was found stable and the null hypothesis of no structural break was not rejected (same applies to “Stabl.” as an output for CUSUMQ). “NA” denotes that a certain test parameter was not applicable.

The results of the additional analyses for the periods 1918-1956 and 1957-1978 also failed to provide clear evidence of association between changes in influenza mortality rates and suicide rates in either short-term or long-term. However, in the analysis among women, a long-term asymmetry was suggested in both periods (the Wald test for asymmetry in the long-term [Wald_LR_] p=0.044 and p<0.001 in the analyses of 1918-1956 and 1957-1978, respectively) and a short-term asymmetry in the analysis of 1957-1978 (Wald_SR_ test p=0.019) (**Tables 2** and **3**). We assume that these findings may again reflect the influence of unobserved confounders, as these results were not supported by other coefficients. It is important to mention that the results for 1957-1978 should be considered with caution since the analysis included time series with only 22 observations. Full specifications for the models used in the analyses of 1918-1956 and 1957-1978 are reported in **Supplementary Table 3** and **4**. No evidence of long-term cointegration between influenza and suicide rates were found in either period (F_PSS statistics does not reject the null hypothesis in the analyses of the whole population, men, and women; **Supplementary Table 3** and **4**, see footnotes for details on bounds test for cointegration).

**Table 2.** Short-term and long-term relationship of suicide rates with positive and negative changes in influenza death rates among the whole population, men, and women in 1918-1956 in Sweden

|  | Whole population |  | Men |  | Women |  |
| --- | --- | --- | --- | --- | --- | --- |
|  | Coeff. (95% CI) | p-value | Coeff. (95% CI) | p-value | Coeff. (95% CI) | p-value |
| **Short-term coefficients** |  |  |  |  |  |  |
| Influenza + | 0.00002<br>(-0.00077 to 0.00082) | 0.955 | -0.00014<br>(-0.00102 to 0.00075) | 0.755 | 0.00098<br>(-0.00068 to 0.00263) | 0.237 |
| Influenza – | 0.00087<br>(-0.00643 to 0.00817) | 0.809 | -0.00132<br>(-0.01001 to 0.00741) | 0.760 | 0.00434<br>(-0.00595 to 0.01463) | 0.394 |
| **Long-term coefficients** |  |  |  |  |  |  |
| Influenza + | 0.00011 | 0.994 | -0.00490 | 0.802 | 0.01403 | 0.521 |
| Influenza – | 0.00103 | 0.936 | 0.00548 | 0.775 | -0.01098 | 0.600 |
| **Model diagnostics** |  |  |  |  |  |  |
| Q-test for autocorrelation, $\chi^2$ | 14.130 | 0.658 | 9.674 | 0.917 | 9.614 | 0.919 |
| Heteroscedasticity, $\chi^2$ | 0.939 | 0.333 | 0.768 | 0.381 | 5.042 | 0.025 |
| Normality, $\chi^2$ | 0.420 | 0.811 | 0.132 | 0.936 | 2.383 | 0.304 |
| RESET, F-statistics | 0.413 | 0.745 | 0.764 | 0.524 | 0.505 | 0.683 |
| CUSUM | Stabl., no str. break | NA | Stabl., no str. break | NA | Stabl., no str. break | NA |
| CUSUMQ | Stabl. | NA | Stabl. | NA | Stabl. | NA |
| Adj. R<sup>2</sup> | 0.309 | NA | 0.345 | NA | 0.432 | NA |
| Wald<sub>SR</sub>, F-statistics | 0.056 | 0.815 | 0.155 | 0.697 | 0.183 | 0.673 |
| Wald<sub>LR</sub>, F-statistics | 1.555 | 0.222 | 0.217 | 0.645 | 4.479 | 0.044 |

*Note:* Signs as “+” and “–” denote the exposure variable being partitioned in positive and negative changes, respectively. Wald test for asymmetry in the short-term (Wald<sub>SR</sub>) and long-term (Wald<sub>LR</sub>) has a null hypothesis of no cointegration. Q-test for autocorrelation reports the results of Portmanteau test for white noise. Heteroscedasticity is measured by Breusch-Pagan/Cook-Weisberg test. Normality is measured by Jarque-Bera test. RESET statistics refers to the regression specification error tests. CUSUM and CUSUMQ refer to the tests of the cumulative sum of recursive residuals and their squares, respectively. “Stabl., no str. break” in the results of CUSUM indicates that the model was found stable and the null hypothesis of no structural break was not rejected (same applies to “Stabl.” as an output for CUSUMQ). “NA” denotes that a certain test parameter was not applicable.

**Table 3.** Short-term and long-term relationship of suicide rates with positive and negative changes in influenza death rates among the whole population, men, and women in 1957-1978 in Sweden

|  | Whole population |  | Men |  | Women |  |
| --- | --- | --- | --- | --- | --- | --- |
|  | Coeff. (95% CI) | p-value | Coeff. (95% CI) | p-value | Coeff. (95% CI) | p-value |
| **Short-term coefficients** |  |  |  |  |  |  |
| Influenza + | -0.00964<br>(-0.02252 to 0.00324) | 0.883 | -0.00876<br>(-0.02738 to 0.00985) | 0.328 | -0.00976<br>(-0.02033 to 0.00081) | 0.068 |
| Influenza – | 0.00194<br>(-0.02614 to 0.03003) | 0.130 | -0.01531<br>(-0.04658 to 0.01596) | 0.309 | 0.02155<br>(-0.00343 to 0.04653) | 0.085 |
| **Long-term coefficients** |  |  |  |  |  |  |
| Influenza + | 0.01255 | 0.618 | -0.02659 | 0.541 | 0.04091 | 0.064 |
| Influenza – | -0.01033 | 0.684 | 0.024538 | 0.582 | -0.02870 | 0.184 |
| **Model diagnostics** |  |  |  |  |  |  |
| Q-test for autocorrelation, $\chi^2$ | 5.662 | 0.773 | 3.790 | 0.925 | 5.243 | 0.813 |
| Heteroscedasticity, $\chi^2$ | 0.102 | 0.750 | 0.155 | 0.694 | 0.079 | 0.778 |
| Normality, $\chi^2$ | 1.569 | 0.456 | 1.627 | 0.443 | 1.237 | 0.537 |
| RESET, F-statistics | 5.839 | 0.014 | 3.348 | 0.064 | 1.089 | 0.398 |
| CUSUM | Stabl., no str. break | NA | Stabl., no str. break | NA | Stabl., no str. break | NA |
| CUSUMQ | Stabl. | NA | Stabl. | NA | Stabl. | NA |
| Adj. R<sup>2</sup> | 0.246 | NA | 0.337 | NA | 0.371 | NA |
| Wald<sub>SR</sub>, F-statistics | 2.903 | 0.112 | 0.536 | 0.477 | 7.132 | 0.019 |
| Wald<sub>LR</sub>, F-statistics | 0.922 | 0.354 | 0.297 | 0.595 | 26.74 | <0.001 |

*Note:* Signs as “+” and “–” denote the exposure variable being partitioned in positive and negative changes, respectively. Wald test for asymmetry in the short-term (Wald<sub>SR</sub>) and long-term (Wald<sub>LR</sub>) has a null hypothesis of no cointegration. Q-test for autocorrelation reports the results of Portmanteau test for white noise. Heteroscedasticity is measured by Breusch-Pagan/Cook-Weisberg test. Normality is measured by Jarque-Bera test. RESET statistics refers to the regression specification error tests. CUSUM and CUSUMQ refer to the tests of the cumulative sum of recursive residuals and their squares, respectively. “Stabl., no str. break” in the results of CUSUM indicates that the model was found stable and the null hypothesis of no structural break was not rejected (same applies to “Stabl.” as an output for CUSUMQ). “NA” denotes that a certain test parameter was not applicable.

A descriptive analysis of mortality data from Statistics Sweden for January-June 2000-2020 (**Figure 3**) showed a considerable fluctuation over time in influenza death rates along with a close correspondence in those rates in men and women. For both sexes, the highest influenza death rates in January-June appeared in 2000 and 2018. During the first six months of 2020, influenza death rates were low in both men and women (1.5 and 1.0 per 100 000 inhabitants, respectively), contrasting the very high COVID-19 death rates observed during the same period (67.8 and 39.9 per 100 000 inhabitants in men and women, respectively). For suicides, data from January-June of each year from 2000 to 2020 revealed, on average, 2.5-time higher rates in men. Interestingly, the relative change in suicide rates in the first half of 2020 compared to suicide rates in the first half of 2019 showed a decrease in rates, although marginal in men (−1.2%; from 8.6 per 100 000 in January-June 2019 to 8.5 per 100 000 in January-June 2020), while more noticeable in women (−12.8%; from 3.9 per 100 000 in January-June 2019 to 3.4 per 100 000 in January-June 2020).

**Figure 3.**
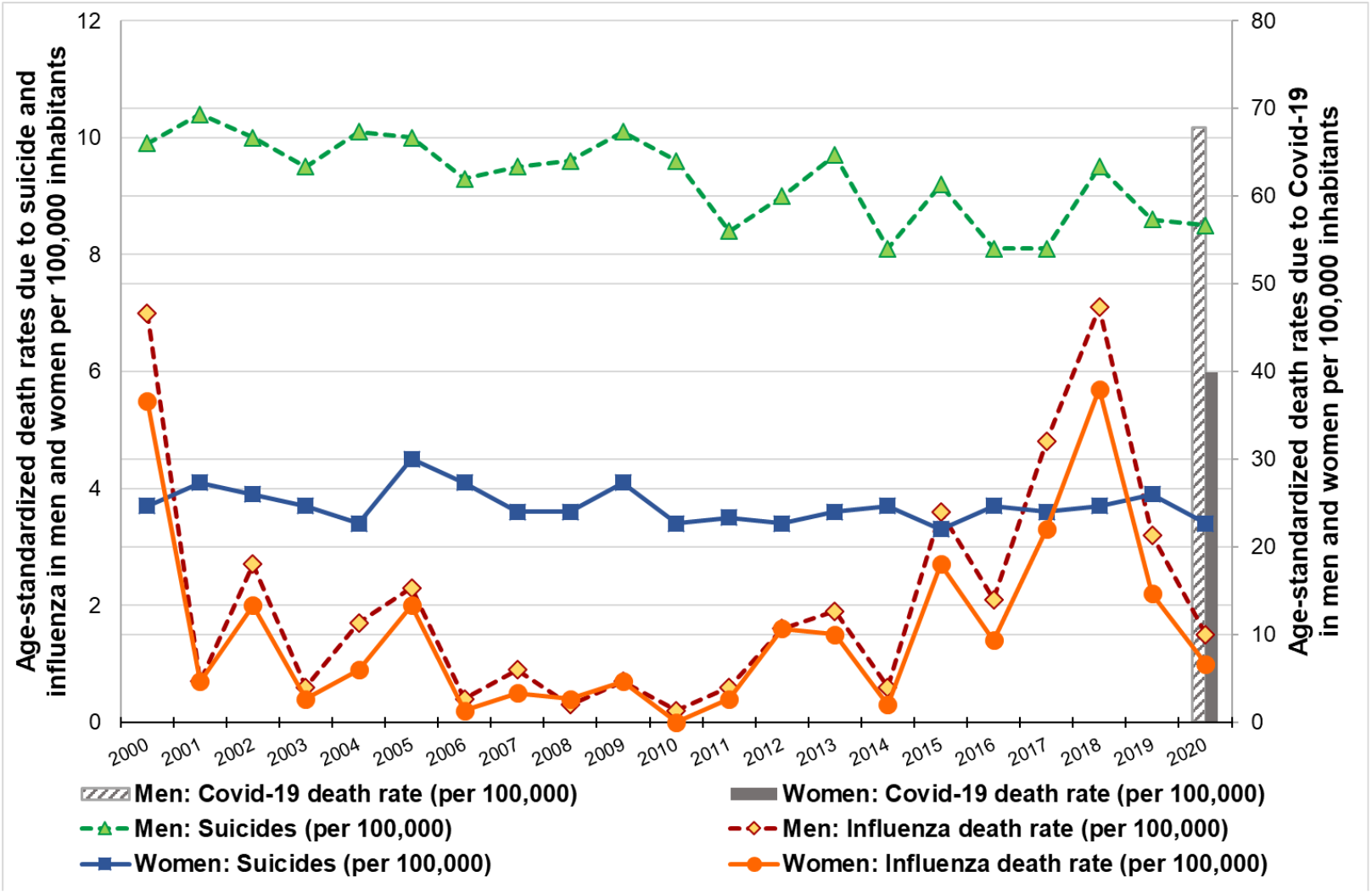
Age-standardized death rates due to suicide and influenza (scaled on the left Y-axis) in January-June 2000-2020 and age-standardized death rates due to Covid-19 (scaled on the right Y-axis) in January-June 2020 in Sweden among men and women per 100 000 inhabitants of corresponding sex. *Note*: Bars denote Covid-19 death rates in first half of 2020. All death rates in the figure are presented on arithmetic scales. The reported death rates are retrieved from Statistics Sweden tables published on November 17, 2020.[25]

## DISCUSSION

This study used publicly available Swedish national data from 1910 to 1978 to shed light on the potential association between influenza-related deaths and over 80 000 deaths by suicide across the three 20^th^ century pandemics. To our knowledge, this is the first study of influenza death and suicide that analyses data from several influenza pandemics. The full modelling provided no clear evidence of either a short-term or a long-term relationship between changes in influenza death rates and changes in suicide rates. The year with the highest number of influenza-related deaths by far, 1918, had the lowest number of suicides in the whole time series. Consistently, preliminary data from the COVID-19 pandemic revealed that suicide rates had slightly decreased in January-June 2020, compared to the rates in January-June 2019 (relative decrease by −1.2% among men and −12.8% among women).

International historical data on pandemics and suicide are very scarce. A report from the US focusing on the years 1910-1920 and using monthly data suggested a possible association between influenza deaths and suicide.[14] Similarly, a study focusing on SARS and suicide found some evidence of sex-specific effects in the short-term.[15] As suicide rates varied considerably in the years preceding SARS in Hong Kong, it is difficult to be certain that SARS, and not other contributing factors, was causally associated with an increase in suicides. In the present study, which uniquely spanned over several decades, we found that while suicide was consistently more prevalent among men throuought the study period, there were no sex-specific associations to influenza death.

What do these results mean for our understanding of the short- and long-term consequences of COVID-19? Our results do not support the widespread belief that global pandemics may necessarily lead to a substantial increase in suicides. The fact that not even the most severe of the 20^th^ century pandemics, the Spanish flu, was associated with increases in suicide in the present study should be kept in mind before making catastrophic predictions of a sharp increase of suicides during and after the current pandemic. There are many factors associated with a global pandemic that may potentially increase suicide risk in the population, but these may be offset by other protective factors that should not be overlooked. A shared sense of belonging and focus, social connectedness and a “pulling together effect” may be one such factor.[1,24]

### Limitations

The study used the Swedish historical public death records and we had no means of verifying the causes of death. The coverage and precision of these records is likely to have improved over time for both variables. To guard against effects of changes in the recording of causes of death, we created a series of dummy variables for each corresponding period but no significant effects of those were found. If there were some other time-varying factors that could affect the coverage and precision of death records, apart from the official changes in registration system, this might have biased our results, in particular if such factors differentially affected the quality of recording deaths due to influenza and suicide. As we did not have access to data with higher temporal resolution than yearly data, that could affect our results if the time sequence of association between changes in influenza death and suicide differ from the chosen time interval. A multitude of factors may vary that impact the resilience of the society with regards to the effect of a pandemic. Such factors may also vary over time and place. However, the fact that we observed no clear associaionts between influenza and suicide deaths across pandemics, which challenged society with various degrees of lockdown, economic effects and health care supply issues, does support the stability of our findings.

## Conclusion

In this national analysis of historical data spanning across three 20^th^ century pandemics, we found no evidence of a short- or long-term association between influenza death rates and suicide rates. Suicide rates in January-June 2020 (when the first wave of COVID-19 occurred) were not higher than those of the corresponding period in 2019. Our results challenge the notion that a tsunami of suicides is to be expected as a result of COVID-19 and we suggest a more moderated message to the public to avoid unfounded fear and an ineffective use of resources for prevention and care.

## Data Availability

Data will be made available upon final publication.

## SUMMARY BOX

### What is already known on this topic

- Little is known on the short- and long-term effects of pandemics on suicide rates.
- Studies of the SARS epidemic suggest sex specific effects.
- Preliminary data from several countries suggest that no rise in suicide rates have followed in the first phase of the COVID-19 pandemic.

### What this study adds

- This study covering the 3 past pandemics in Sweden do not provide evidence of significant associations between influenza death rates and changes in suicide rates.
- Suicide rates in January-June 2020, during the first wave of COVID-19, revealed a slight decrease compared to the corresponding rates in January-June 2019.
- These findings should help policymakers and clinicians to prioritize how to use resources to best counter the effects of the current and future pandemics.

### Data sharing statement

The full dataset (numbers of deaths by influenza and suicide and total population for 1910-1978) will be made publicly available at OSF.io with unrestricted access. Data on deaths in January-June 1997-2020 are available online at the National Board of Health and Welfare.[20,25]

### Transparency declaration

The lead author affirms that this manuscript is an honest, accurate, and transparent account of the study being reported; that no important aspects of the study have been omitted; and that any discrepancies from the study as planned have been explained.

