## Supplementary material for "Will the COVID-19 pandemic lead to a tsunami of suicides? A Swedish nationwide analysis of historical and 2020 data"

**Model diagnostics**

We carried out several types of model diagnostics, including Portmanteau test for white noise to check autocorrelation in residuals, Breusch-Pagan/Cook-Weisberg test for heteroscedasticity, Jarque-Bera test on normality, and the regression specification error tests that indicates whether there is a misspecification in the model. We also performed tests for stability of the models by using the cumulative sum of recursive residuals and their squares to test whether there is a structural break due to changes in regression coefficients over time.

**STATA codes for the non-linear autoregressive distributed lag (NARDL) modelling of association between between influenza death rates and suicide rates**

```
clear
use "Z:\Influenza_deaths_suicides\ALL19101978.dta"


///// 1. DATA MANAGEMENT: generate and label variables to be used in the analyses


/// Labels for original variables (retrieved from the Statistical Yearbooks 1910-1978)

 lab var year "Years 1910-1978"

 lab var number_population_all "Total number of inhabitants (population size) in Sweden in corresponding year"
 lab var number_population_men "Total number of male population in Sweden in corresponding year"
 lab var number_population_women "Total number of female population in Sweden in corresponding year"

 lab var number_influenza_all "Number of deaths from influenza in total population in corresponding year"
 lab var number_influenza_men "Number of deaths from influenza among men in corresponding year"
 lab var number_influenza_women "Number of deaths from influenza among women in corresponding year"

 lab var number_suicide_all "Number of suicides in total population in corresponding year"
 lab var number_suicide_men "Number of suicides among men in corresponding year"
 lab var number_suicide_women "Number of suicides among women in corresponding year"



/// Calculation of annual influenza death rates and suicide rates per 100,000 for total population, men, and
women

 gen influenza_rates_all = (number_influenza_all/number_population_all)*100000
 gen influenza_rates_men = (number_influenza_men/number_population_men)*100000
 gen influenza_rates_women = (number_influenza_women/number_population_women)*100000
```

/// Logarithmic transformation for suicide rates for total population, men, and women

```
gen ln_suicide_rates_all = ln(suicide_rates_all)
gen ln_suicide_rates_men = ln(suicide_rates_men)
gen ln_suicide_rates_women = ln(suicide_rates_women)

lab var ln_suicide_rates_all "Log-transformed suicide rates for total population per 100,000, annual"
lab var ln_suicide_rates_men "Log-transformed suicide rates among men per 100,000, annual"
lab var ln_suicide_rates_women "Log-transformed suicide rates among women per 100,000, annual"
```

/// Variables for changes in death registration in Sweden 1910-1978 (as dummy variables)

```
// based on the Bertillon criteria (prior to 1931)
gen registration_19101930=0
replace registration_19101930=1 if year<=1930
lab var registration_19101930 "Dummy variable for death registration in 1910-1930 (Bertillion)"

// introduced in cooperation with other Nordic countries (1931-1950)
gen registration_19311950=0
replace registration_19311950=1 if year>=1931 & year<=1950
lab var registration_19311950 "Dummy variable for death registration in 1931-1950 (new registration)"

// ICD-6 (1951-1957)
gen registration_19511957=0
replace registration_19511957=1 if year>=1951 & year<=1957
lab var registration_19511957 "Dummy variable for death registration in 1951-1957 (ICD-6)"

// ICD-7 (1958-1968)
gen registration_19581968=0
replace registration_19581968=1 if year>=1958 & year<=1968
lab var registration_19581968 "Dummy variable for death registration in 1958-1968 (ICD-7)"

// ICD-8 (1969-1978)
gen registration_1969after=0
replace registration_1969after=1 if year>=1969
lab var registration_1969after "Dummy variable for death registration in 1969 and after (ICD-8)"
////////////////////////////////////////////////////////////////////////////
```

///// 2. CHECKING VARIABLES' PROPERTIES AND TESTING THE CONDITIONS FOR MODELLING

/// Declare data to be time-series data

```
tsset year, yearly
```

/// Obtain optimal lags for each variable (for (i) augmentation in ADF and KPSS tests, and (ii) for p and q parameters in NARDL).

/// Lags obtained for influenza death rates and logarithmically-transformed suicide rates for total population, men, and women, and for time periods 1910-1978, 1918-1956, and 1957-1978

/// If AIC, HQIC, and SBIC information criteria indicated different lag orders, SBIC was used to select optimal lags

```
varsoc influenza_rates_all
varsoc influenza_rates_all if tin(1918, 1956)
```

```
varsoc influenza_rates_all if tin(1957, )

varsoc influenza_rates_men
varsoc influenza_rates_men if tin(1918, 1956)
varsoc influenza_rates_men if tin(1957, )

varsoc influenza_rates_women
varsoc influenza_rates_women if tin(1918, 1956)
varsoc influenza_rates_women if tin(1957, )

varsoc ln_suicide_rates_all
varsoc ln_suicide_rates_all if tin(1918, 1956)
varsoc ln_suicide_rates_all if tin(1957, )

varsoc ln_suicide_rates_men
varsoc ln_suicide_rates_men if tin(1918, 1956)
varsoc ln_suicide_rates_men if tin(1957, )

varsoc ln_suicide_rates_women
varsoc ln_suicide_rates_women if tin(1918, 1956)
varsoc ln_suicide_rates_women if tin(1957, )


/// Tests for stationarity: ADF and KPSS for influenza death rates and logarithmically-transformed suicide rates variables for total population, men, and women
/// Augmentation by at least one lag was used (for suicide rates in women - by two lags according to SBIC in varsoc)

dfuller influenza_rates_all, lag(1)
dfuller influenza_rates_men, lag(1)
dfuller influenza_rates_women, lag(1)

dfuller ln_suicide_rates_all, lag(1)
dfuller ln_suicide_rates_men, lag(1)
dfuller ln_suicide_rates_women, lag(1)
dfuller ln_suicide_rates_women, lag(2)

dfuller D.influenza_rates_all, lag(1)
dfuller D.influenza_rates_men, lag(1)
dfuller D.influenza_rates_women, lag(1)

dfuller D.ln_suicide_rates_all, lag(1)
dfuller D.ln_suicide_rates_men, lag(1)
dfuller D.ln_suicide_rates_women, lag(1)
dfuller D.ln_suicide_rates_women, lag(2)

kpss influenza_rates_all, maxlag(1) notrend
kpss influenza_rates_men, maxlag(1) notrend
kpss influenza_rates_women, maxlag(1) notrend

kpss ln_suicide_rates_all, maxlag(1) notrend
kpss ln_suicide_rates_men, maxlag(1) notrend
kpss ln_suicide_rates_women, maxlag(2) notrend

kpss D.influenza_rates_all, maxlag(1) notrend
kpss D.influenza_rates_men, maxlag(1) notrend
kpss D.influenza_rates_women, maxlag(1) notrend

kpss D.ln_suicide_rates_all, maxlag(1) notrend
kpss D.ln_suicide_rates_men, maxlag(1) notrend
```

kpss D.ln_suicide_rates_women, maxlag(2) notrend

///// 3. ESTIMATION OF NON-LINEAR AUTOREGRESSIVE DISTRIBUTED LAG (NARDL) MODELS: for total population, men, and women, and for time periods 1910-1978, 1918-1956, and 1957-1978
/// The dependent and independent variables are indicated in levels.
/// The covariates (i.e., changes in death registration system 1910-1978) are included with the deterministic option, but these were only kept in the models if statistically significant.
/// The optimal number of lags for dependent and independent variables (p and q parameters, respectively).
/// Since p and q parameters refer to levels, one additional lag is added to p and q to get an optimal lag length in differences (p and q must be at least 2).
/// The model provides an output for a long-term cointegration bounds test and diagnostic tests.

nardl ln_suicide_rates_all influenza_rates_all, p(2) q(2) h(69) plot bootstrap(100) level(95) residuals /* used as a final model */
nardl ln_suicide_rates_all influenza_rates_all, p(2) q(2) deterministic(registration_19101930 registration_19311950 registration_19511957 registration_19581968 registration_1969after) h(69) plot bootstrap(100) level(95) residuals

nardl ln_suicide_rates_men influenza_rates_men, p(2) q(2) h(69) plot bootstrap(100) level(95) residuals /* used as a final model */
nardl ln_suicide_rates_men influenza_rates_men, p(2) q(2) deterministic(registration_19101930 registration_19311950 registration_19511957 registration_19581968 registration_1969after) h(69) plot bootstrap(100) level(95) residuals

nardl ln_suicide_rates_women influenza_rates_women, p(3) q(2) h(69) plot bootstrap(100) level(95) residuals /* used as a final model */
nardl ln_suicide_rates_women influenza_rates_women, p(3) q(2) deterministic(registration_19101930 registration_19311950 registration_19511957 registration_19581968 registration_1969after) h(69) plot bootstrap(100) level(95) residuals

nardl ln_suicide_rates_all influenza_rates_all if tin(1918,1956), p(2) q(2) h(39) plot bootstrap(50) level(95) residuals
nardl ln_suicide_rates_men influenza_rates_men if tin(1918,1956), p(2) q(2) h(39) plot bootstrap(50) level(95) residuals
nardl ln_suicide_rates_women influenza_rates_women if tin(1918,1956), p(5) q(2) h(39) plot bootstrap(50) level(95) residuals

nardl ln_suicide_rates_all influenza_rates_all if tin(1957, ), p(2) q(2) h(22) plot bootstrap(50) level(95) residuals
nardl ln_suicide_rates_men influenza_rates_men if tin(1957, ), p(2) q(2) h(22) plot bootstrap(50) level(95) residuals
nardl ln_suicide_rates_women influenza_rates_women if tin(1957, ), p(2) q(2) h(22) plot bootstrap(50) level(95) residuals

/* REFERENCES
For varsoc and dfuller:
StataCorp. Stata 15 Base Reference Manual. College Station, TX: Stata Press; 2017

For kpss:
Author: Christopher F Baum, Boston College, USA baum@@bc.edu
Kwiatkowski, D., Phillips, P.C.B., Schmidt, P. and Y. Shin. Testing the null hypothesis of stationarity against the alternative of a unit root: How sure are we that economic time series have a unit root? Journal of Econometrics, 54, 1992, 159-178.
Lee, D. and P. Schmidt. On the power of the KPSS test of stationarity against fractionally-integrated alternatives. Journal of Econometrics, 73, 1996, 285-302.
Schwert, G.W. Tests for Unit Roots: A Monte Carlo Investigation. Journal of Business and Economic Statistics, 7, 1989, 147-160.

For nardl:
Author: Stata implementation by Marco Sunder. This version was created on 26jan2012.
Shin, Y., Yu, B., Greenwood-Nimmo, M. (2011): Modelling asymmetric cointegration and dynamic multipliers in a nonlinear ARDL framework. Working paper (version of November 2011), http://ssrn.com/abstract=1807745. */

end of do-file

**Supplementary Table 1. Unit root tests at the level and first difference by Augmented Dickey-Fuller (ADF) and Kwiatkowski-Phillips-Schmidt-Shin (KPSS) test statistics**

| | ADF at level | | KPSS at level | | Results for stationarity at the level | ADF at first difference | | KPSS at first difference | | Results for stationarity at the 1st difference | Order of integration |
|---|---|---|---|---|---|---|---|---|---|---|---|
| | Test statistic | p-value | Test statistic | p-value[1] | | Test statistic | p-value | Test statistic | p-value[1] | | |
| **Suicide rate (log-transformed)** | | | | | | | | | | | |
| All | -2.181 | 0.213 | 1.980 | <0.001 | nonstationary | -6.640 | <0.001 | 0.068 | >0.1 | stationary | 1 |
| Males | -3.230 | 0.018 | 0.964 | <0.001 | nonstationary | -6.512 | <0.001 | 0.053 | >0.1 | stationary | 1 |
| Females | -0.517 | 0.889 | 3.130 | <0.001 | nonstationary | -8.692 | <0.001 | 0.104 | >0.1 | stationary | 1 |
| **Influenza death rate** | | | | | | | | | | | |
| All | -4.895 | <0.001 | 0.461 | >0.1 | stationary | *-8.727* | *<0.001* | *0.016* | >0.1 | *stationary* | 0 |
| Males | -4.856 | <0.001 | 0.431 | >0.1 | stationary | *-8.642* | *<0.001* | *0.016* | >0.1 | *stationary* | 0 |
| Females | -4.940 | <0.001 | 0.491 | >0.1 | stationary | *-8.824* | *<0.001* | *0.015* | >0.1 | *stationary* | 0 |

*Note*: For the ADF test, the null hypothesis implies that the variable contains a unit root (the alternate hypothesis is that the variable is stationary), whereas for the KPSS test the null hypothesis implies that the variable is stationary (the alternate hypothesis is that there is a unit root). The results for ADF and KPSS tests for stationarity at first difference for influenza death rates are reported as explanatory (written in Italics) since the stationarity at the level has already been established (i.e., integrated of the order zero).

[1] KPSS test results do not indicate the exact p-value, but report the level of significance at which the null hypothesis is rejected (1%, 2.5%, 5%, or 10% significance level).

Abbreviations: ADF, Augmented Dickey–Fuller unit-root test; KPSS, Kwiatkowski-Phillips-Schmidt-Shin test for stationarity

**Supplementary Table 2. Full specification for non-linear autoregressive distributed lag models used for the analysis of short-term and long-term relationship of suicide rates with positive and negative changes in influenza death rates among the whole population, men, and women in 1910-1978 in Sweden**

| | Whole population | | Men | | Women | |
|---|---|---|---|---|---|---|
| | **Coefficient (95% CI)** | **p-value** | **Coefficient (95% CI)** | **p-value** | **Coefficient (95% CI)** | **p-value** |
| LnSuicide, lag 1 | -0.08466 (-0.23940 to 0.07009) | 0.278 | -0.11457 (-0.30588 to 0.07674) | 0.235 | -0.04517 (-0.15832 to 0.06799) | 0.427 |
| Influenza +, lag 1 | -0.00001 (-0.00761 to 0.00759) | 0.998 | -0.00150 (-0.01032 to 0.00732) | 0.734 | 0.00508 (-0.00366 to 0.01382) | 0.249 |
| Influenza –, lag 1 | -0.00018 (-0.00775 to 0.00739) | 0.962 | -0.00165 (-0.01044 to 0.00714) | 0.708 | 0.00487 (-0.00383 to 0.01358) | 0.267 |
| ΔLnSuicide, lag 1 | -0.14610 (-0.43113 to 0.13893) | 0.309 | 0.029465 (-0.27204 to 0.33097) | 0.846 | -0.65184 (-0.91785 to -0.38582) | <0.001 |
| ΔLnSuicide, lag 2 | NA | NA | NA | NA | -0.30807 (-0.55722 to -0.05892) | 0.016 |
| ΔInfluenza + | 0.00002 (-0.00036 to 0.00039) | 0.931 | 0.00004 (-0.00034 to 0.00041) | 0.854 | -0.00007 (-0.00056 to 0.00041) | 0.760 |
| ΔInfluenza +, lag 1 | 0.00153 (-0.00285 to 0.00590) | 0.488 | 0.00085 (-0.00347 to 0.00518) | 0.694 | 0.00093 (-0.00482 to 0.00668) | 0.747 |
| ΔInfluenza – | 0.00103 (-0.00579 to 0.00785) | 0.764 | -0.00192 (-0.01005 to 0.00621) | 0.638 | 0.00780 (0.00015 to 0.01544) | 0.046 |
| ΔInfluenza –, lag 1 | -0.00070 (-0.00236 to 0.00096) | 0.401 | -0.00038 (-0.00212 to 0.00136) | 0.663 | -0.000075 (-0.00289 to 0.00139) | 0.488 |
| **Long-term effect** | | | | | | |
| Influenza + | -0.00012 | 0.998 | -0.01314 | 0.745 | 0.11254 | 0.538 |
| Influenza – | 0.00211 | 0.962 | 0.01443 | 0.722 | -0.10789 | 0.544 |

| | | | | | | |
|---|---|---|---|---|---|---|
| **Model diagnostics** | | | | | | |
| Q-test for autocorrelation, $\chi^2$ | 40.160 | 0.125 | 35.260 | 0.273 | 31.850 | 0.424 |
| Heteroscedasticity, $\chi^2$ | 3.649 | 0.056 | 5.439 | 0.020 | 1.280 | 0.258 |
| Normality, $\chi^2$ | 2.826 | 0.243 | 2.237 | 0.327 | 2.372 | 0.305 |
| RESET, F-statistics | 4.656 | 0.006 | 5.473 | 0.023 | 1.013 | 0.394 |
| CUSUM | Stable, no structural break | NA | Stable, no structural break | NA | Stable, no structural break | NA |
| CUSUMQ | Stable | NA | Stable | NA | Stable | NA |
| Adj. $R^2$ | 0.257 | NA | 0.300 | NA | 0.374 | NA |
| $Wald_{SR}$, F-statistics | 0.054 | 0.817 | 0.275 | 0.602 | 1.062 | 0.307 |
| $Wald_{LR}$, F-statistics | 1.206 | 0.277 | 1.176 | 0.283 | 0.771 | 0.384 |
| **Cointegration test statistics** | | | | | | |
| t_BDM | -1.095 | NA | -1.199 | NA | -0.799 | NA |
| F_PSS | 3.789 | NA | 3.735 | NA | 2.530 | NA |
| **Critical values for F_PSS** | | | | | | |
| 5% critical values; I(0), I(1) | k=1: 5.055, 5.915<br>k=2: 3.947, 5.020 | NA | k=1: 5.055, 5.915<br>k=2: 3.947, 5.020 | NA | k=1: 5.055, 5.915<br>k=2: 3.947, 5.020 | NA |

*Note*: F_PSS denotes F-statistics for Pesaran/Shin/Smith bounds test with null hypothesis of no cointegration. Critical values are retrieved from Narayan PK (2005) for a sample size of n=70.[1] The null hypothesis of no cointegration is supported if F_PSS statistics is lower than 5% I(0) critical value; the null hypothesis is rejected if F_PSS statistics is greater than 5% I(1) critical value. We applied a conservative approach to considering the number of exposure variables (k), as suggested by Shin Y, et al (2014) [2] (since influenza death rates represent one exposure (k=1), while the analysis actually partitions the exposure to positive and negative changes (k=2)). In a given table, the results of F_PSS for the whole population, men, and women are lower than the reported 5% I(0) critical values that accepts a null hypothesis of no cointegration.

"LnSuicide" indicates that suicide rates were logarithmically-transformed. "Δ" signifies the series is differenced. Signs as "+" and "–" denote the exposure variable being partitioned in positive and negative changes, respectively. "NA" denotes that a certain test or test parameter is not applicable.

Number of lags used for each variable in the model are noted by "lag#". To select the optimal number of lags to be used for choosing p and q parameters for the NARDL (i.e., numbers of lags for dependent and independent variables, respectively), we applied a varsoc command in STATA using the minimal values of Akaike Information Criterion, Schwarz's Bayesian information criterion (SBIC), and the Hannan and Quinn information criterion information criteria. If information criteria indicated different lag orders, SBIC was used to select optimal lags. Thus, for the analysis of the whole population and men, both exposure and outcome time series were lagged once, while for analysis among women to outcome time series were lagged twice and exposure time series were lagged once.

Wald test for asymmetry in a short-term ($Wald_{SR}$) and long-term ($Wald_{LR}$) has a null hypothesis of no cointegration. Q-test for autocorrelation reports the results of Portmanteau test for white noise. Heteroscedasticity is measured by Breusch-Pagan/Cook-Weisberg test. Normality is measured by Jarque-Bera test. RESET statistics refers to the regression specification error tests. CUSUM and CUSUMQ refer to the tests of the cumulative sum of recursive residuals and their squares, respectively. "Stable, no structural break" in the results of CUSUM indicates that the model was found stable and the null hypothesis of no structural break was not rejected (same applies to "Stable" as an output for CUSUMQ).

**Supplementary Table 3. Full specification for non-linear autoregressive distributed lag models used for the analysis of short-term and long-term relationship of suicide rates with positive and negative changes in influenza death rates among the whole population, men, and women in 1918-1956 in Sweden**

| | Whole population | | Men | | Women | |
|---|---|---|---|---|---|---|
| | Coefficient (95% CI) | p-value | Coefficient (95% CI) | p-value | Coefficient (95% CI) | p-value |
| LnSuicide, lag 1 | -0.32229 (-0.73472 to 0.09015) | 0.121 | -0.26417 (-0.60333 to 0.07492) | 0.122 | -0.36067 (-0.97384 to 0.25250) | 0.238 |
| Influenza +, lag 1 | 0.00003 (-0.00851 to 0.00858) | 0.994 | -0.00129 (-0.011384 to 0.00879) | 0.795 | 0.00506 (-0.00682 to 0.01695) | 0.390 |
| Influenza –, lag 1 | -0.00033 (-0.00869 to 0.00803) | 0.936 | -0.00145 (-0.01129 to 0.00840) | 0.766 | 0.00396 (-0.00801 to 0.01593) | 0.503 |
| ΔLnSuicide, lag 1 | -0.19387 (-0.59955 to 0.21181) | 0.337 | -0.03045 (-0.44243 to 0.38153) | 0.881 | -0.58189 (-1.19784 to 0.03406) | 0.063 |
| ΔLnSuicide, lag 2 | NA | NA | NA | NA | -0.36148 (-0.91643 to 0.19346) | 0.193 |
| ΔLnSuicide, lag 3 | NA | NA | NA | NA | -0.11520 (-0.61658 to 0.38617) | 0.641 |
| ΔLnSuicide, lag 4 | NA | NA | NA | NA | 0.11306 (-0.26409 to 0.49021) | 0.544 |
| ΔInfluenza + | 0.00002 (-0.00077 to 0.00082) | 0.955 | -0.00014 (-0.00102 to 0.00075) | 0.755 | 0.00098 (-0.00068 to 0.00263) | 0.237 |
| ΔInfluenza +, lag 1 | 0.00140 (-0.00347 to 0.00627) | 0.561 | 0.00095 (-0.00389 to 0.00580) | 0.691 | -0.00063 (-0.00821 to 0.00696) | 0.866 |
| ΔInfluenza – | 0.00087 (-0.00643 to 0.00817) | 0.809 | -0.00132 (-0.01001 to 0.00741) | 0.760 | 0.00434 (-0.00595 to 0.01463) | 0.394 |
| ΔInfluenza –, lag 1 | -0.00076 (-0.00256 to 0.00104) | 0.396 | -0.00044 (-0.00233 to 0.00145) | 0.637 | -0.00052 (-0.00328 to 0.00224) | 0.704 |
| **Long-term effect** | | | | | | |

| | | | | | | |
|---|---|---|---|---|---|---|
| Influenza + | 0.00011 | 0.994 | -0.00490 | 0.802 | 0.01403 | 0.521 |
| Influenza – | 0.00103 | 0.936 | 0.00548 | 0.775 | -0.01098 | 0.600 |
| **Model diagnostics** | | | | | | |
| Q-test for autocorrelation, $\chi^2$ | 14.130 | 0.658 | 9.674 | 0.917 | 9.614 | 0.919 |
| Heteroscedasticity, $\chi^2$ | 0.939 | 0.333 | 0.768 | 0.381 | 5.042 | 0.025 |
| Normality, $\chi^2$ | 0.420 | 0.811 | 0.132 | 0.936 | 2.383 | 0.304 |
| RESET, F-statistics | 0.413 | 0.745 | 0.764 | 0.524 | 0.505 | 0.683 |
| CUSUM | Stable, no structural break | NA | Stable, no structural break | NA | Stable, no structural break | NA |
| CUSUMQ | Stable | NA | Stable | NA | Stable | NA |
| Adj. $R^2$ | 0.309 | NA | 0.345 | NA | 0.432 | NA |
| $Wald_{SR}$, F-statistics | 0.056 | 0.815 | 0.155 | 0.697 | 0.183 | 0.673 |
| $Wald_{LR}$, F-statistics | 1.555 | 0.222 | 0.217 | 0.645 | 4.479 | 0.044 |
| **Cointegration test statistics** | | | | | | |
| t_BDM | -1.596 | NA | -1.591 | NA | -1.207 | NA |
| F_PSS | 1.009 | NA | 1.019 | NA | 1.328 | NA |
| **Critical values for F_PSS** | | | | | | |

| | | | | | | |
|---|---|---|---|---|---|---|
| 5% critical values; I(0) to I(1) | k=1: 5.260, 6.160<br>k=2: 4.133, 5.260 | NA | k=1: 5.260, 6.160<br>k=2: 4.133, 5.260 | NA | k=1: 5.260, 6.160<br>k=2: 4.133, 5.260 | NA |

*Note*: F_PSS denotes F-statistics for Pesaran/Shin/Smith bounds test with null hypothesis of no cointegration. Critical values are retrieved from Narayan PK (2005) for a sample size of n=40 [1]. The null hypothesis of no cointegration is supported if F_PSS statistics is lower than 5% I(0) critical value; the null hypothesis is rejected if F_PSS statistics is greater than 5% I(1) critical value. We applied a conservative approach to considering the number of exposure variables (k), as suggested by Shin Y, et al (2014) [2] (since influenza death rates represent one exposure (k=1), while the analysis actually partitions the exposure to positive and negative changes (k=2)). In a given table, the results of F_PSS for the whole population, men, and women are lower than the reported 5% I(0) critical values that accepts a null hypothesis of no cointegration.

"LnSuicide" indicates that suicide rates were logarithmically-transformed. "Δ" signifies the series is differenced. Signs as "+" and "–" denote the exposure variable being partitioned in positive and negative changes, respectively. "NA" denotes that a certain test or test parameter is not applicable.

Number of lags used for each variable in the model are noted by "lag#". To select the optimal number of lags to be used for choosing p and q parameters for the NARDL (i.e., numbers of lags for dependent and independent variables, respectively), we applied a varsoc command in STATA using the minimal values of Akaike Information Criterion, Schwarz's Bayesian information criterion (SBIC), and the Hannan and Quinn information criterion information criteria. If information criteria indicated different lag orders, SBIC was used to select optimal lags. Thus, for the analysis of the whole population and men, both exposure and outcome time series were lagged once, while for analysis among women, outcome time series were lagged four times and exposure time series were lagged once.

Wald test for asymmetry in a short-term ($Wald_{SR}$) and long-term ($Wald_{LR}$) has a null hypothesis of no cointegration. Q-test for autocorrelation reports the results of Portmanteau test for white noise. Heteroscedasticity is measured by Breusch-Pagan/Cook-Weisberg test. Normality is measured by Jarque-Bera test. RESET statistics refers to the regression specification error tests. CUSUM and CUSUMQ refer to the tests of the cumulative sum of recursive residuals and their squares, respectively. "Stable, no structural break" in the results of CUSUM indicates that the model was found stable and the null hypothesis of no structural break was not rejected (same applies to "Stable" as an output for CUSUMQ).

**Supplementary Table 4. Full specification for non-linear autoregressive distributed lag models used for the analysis of short-term and long-term relationship of suicide rates with positive and negative changes in influenza death rates among the whole population, men, and women in 1957-1978 in Sweden**

| | Whole population | | Men | | Women | |
|---|---|---|---|---|---|---|
| | **Coefficient (95% CI)** | **p-value** | **Coefficient (95% CI)** | **p-value** | **Coefficient (95% CI)** | **p-value** |
| LnSuicide, lag 1 | -0.64009 (-1.15104 to -0.12914) | 0.018 | -0.52238 (-1.11901 to 0.07425) | 0.081 | -0.66040 (-1.04522 to -0.27558) | 0.003 |
| Influenza +, lag 1 | 0.008034 (-0.02823 to 0.04429) | 0.640 | -0.01385 (-0.05324 to 0.02553) | 0.461 | 0.02702 (-0.00364 to 0.05768) | 0.079 |
| Influenza –, lag 1 | 0.00661 (-0.02823 to 0.04429) | 0.699 | -0.01281 (-0.054045 to 0.02841) | 0.514 | 0.01895 (-0.01066 to 0.04857) | 0.190 |
| ΔLnSuicide, lag 1 | 0.14293 (-0.41621 to 0.70206) | 0.590 | -0.04565 (-0.70111 to 0.60981) | 0.883 | 0.16936 (-0.25995 to 0.59868) | 0.409 |
| ΔInfluenza + | -0.00964 (-0.02252 to 0.00324) | 0.883 | -0.00876 (-0.02738 to 0.00985) | 0.328 | -0.00976 (-0.02033 to 0.00081) | 0.068 |
| ΔInfluenza +, lag 1 | -0.01999 (-0.04573 to 0.00574) | 0.117 | -0.01837 (-0.04871 to 0.01197) | 0.214 | -0.01298 (-0.04073 to 0.014772) | 0.331 |
| ΔInfluenza – | 0.00194 (-0.02614 to 0.03003) | 0.130 | -0.01531 (-0.04658 to 0.01596) | 0.309 | 0.02155 (-0.00343 to 0.04653) | 0.085 |
| ΔInfluenza –, lag 1 | 0.00864 (-0.00271 to 0.02000) | 0.124 | 0.01033 (-0.00294 to 0.02359) | 0.116 | 0.00585 (-0.00565 to 0.01735) | 0.292 |
| **Long-term effect** | | | | | | |
| Influenza + | 0.01255 | 0.618 | -0.02659 | 0.541 | 0.04091 | 0.064 |
| Influenza – | -0.01033 | 0.684 | 0.024538 | 0.582 | -0.02870 | 0.184 |
| **Model diagnostics** | | | | | | |

| | | | | | | |
|---|---|---|---|---|---|---|
| Q-test for autocorrelation, $\chi^2$ | 5.662 | 0.773 | 3.790 | 0.925 | 5.243 | 0.813 |
| Heteroscedasticity, $\chi^2$ | 0.102 | 0.750 | 0.155 | 0.694 | 0.079 | 0.778 |
| Normality, $\chi^2$ | 1.569 | 0.456 | 1.627 | 0.443 | 1.237 | 0.537 |
| RESET, F-statistics | 5.839 | 0.014 | 3.348 | 0.064 | 1.089 | 0.398 |
| CUSUM | Stable, no structural break | NA | Stable, no structural break | NA | Stable, no structural break | NA |
| CUSUMQ | Stable | NA | Stable | NA | Stable | NA |
| Adj. $R^2$ | 0.246 | NA | 0.337 | NA | 0.371 | NA |
| $Wald_{SR}$, F-statistics | 2.903 | 0.112 | 0.536 | 0.477 | 7.132 | 0.019 |
| $Wald_{LR}$, F-statistics | 0.922 | 0.354 | 0.297 | 0.595 | 26.74 | <0.001 |
| **Cointegration test statistics** | | | | | | |
| t_BDM | -2.706 | NA | -1.891 | NA | -3.707 | NA |
| F_PSS | 2.649 | NA | 2.075 | NA | 4.688 | NA |
| **Critical values for F_PSS** | | | | | | |
| 5% critical values; I(0), I(1) | k=1: 5.395, 6.350<br>k=2: 4.267, 5.473 | NA | k=1: 5.395, 6.350<br>k=2: 4.267, 5.473 | NA | k=1: 5.395, 6.350<br>k=2: 4.267, 5.473 | NA |

*Note*: F_PSS denotes F-statistics for Pesaran/Shin/Smith bounds test with null hypothesis of no cointegration. Critical values are retrieved from Narayan PK (2005) for a sample size of n=30 [1]. The null hypothesis of no cointegration is supported if F_PSS statistics is lower than 5% I(0) critical value; the null hypothesis is rejected if F_PSS statistics is greater than 5% I(1) critical value. We applied a conservative approach to considering the number of exposure variables (k), as suggested by Shin Y, et al (2014) [2] (since influenza death rates represent one exposure (k=1), while the analysis actually partitions the exposure to positive and negative changes (k=2)). In a given table, the results of F_PSS for the whole population, men, and women are lower than the reported 5% I(0) critical values that accepts a null hypothesis of no cointegration.

"LnSuicide" indicates that suicide rates were logarithmically-transformed. "Δ" signifies the series is differenced. Signs as "+" and "–" denote the exposure variable being partitioned in positive and negative changes, respectively. "NA" denotes that a certain test or test parameter is not applicable.

Number of lags used for each variable in the model are noted by "lag#". To select the optimal number of lags to be used for choosing p and q parameters for the NARDL (i.e., numbers of lags for dependent and independent variables, respectively), we applied a varsoc command in STATA using the minimal values of Akaike Information Criterion, Schwarz's Bayesian information criterion (SBIC), and the Hannan and Quinn information criterion information criteria. If information criteria indicated different lag orders, SBIC was used to select optimal lags. Thus, for the analysis of the whole population, men, and women, both exposure and outcome time series were lagged once.

Wald test for asymmetry in a short-term ($Wald_{SR}$) and long-term ($Wald_{LR}$) has a null hypothesis of no cointegration. Q-test for autocorrelation reports the results of Portmanteau test for white noise. Heteroscedasticity is measured by Breusch-Pagan/Cook-Weisberg test. Normality is measured by Jarque-Bera test. RESET statistics refers to the regression specification error tests. CUSUM and CUSUMQ refer to the tests of the cumulative sum of recursive residuals and their squares, respectively. "Stable, no structural break" in the results of CUSUM indicates that the model was found stable and the null hypothesis of no structural break was not rejected (same applies to "Stable" as an output for CUSUMQ).
